# Distinct proteomic signatures in Ethiopians predict acute and long-term sequelae of COVID-19

**DOI:** 10.1101/2024.10.09.24315196

**Authors:** Dawit Wolday, Abrha G. Gebrehiwot, Muhammed Ahmed Rameto, Saro Abdella, Atsbeha Gebreegziabxier, Wondwossen Amogne, Tobias F. Rinke de Wit, Messay Hailu, Getachew Tollera, Geremew Tasew, Masresha Tessema, Matthew Miller, Manali Mukhreji, Amy Gillgrass, Dawn M.E. Bowdish, Charu Kaushic, Chris P. Verschoor

**Affiliations:** Department of Biochemistry and Biomedical Sciences, Faculty of Health Sciences, McMaster University, Hamilton, Ontario, Canada; McMaster Immunology Research Centre, Faculty of Health Sciences, McMaster University, Hamilton, Ontario, Canada; Michael G. DeGroote Institute for Infectious Diseases, McMaster University, Hamilton, Ontario, Canada; Ethiopian Public Health Institute, Addis Ababa, Ethiopia; Department of Infectious Diseases, College of Health Sciences, Addis Ababa University, Addis Ababa, Ethiopia; Amsterdam Institute for Global Health and Development, Academic Medical Center – Amsterdam University, Amsterdam, The Netherlands; Department of Medicine, Faculty of Health Sciences, McMaster University, Hamilton, Ontario, Canada; Firestone Institute for Respiratory Health, The Research Institute of St Joe’s, St Joseph’s Healthcare Hamilton, Hamilton, Ontario, Canada; Health Sciences North Research Institute, Northern Ontario School of Medicine University, Sudbury, Ontario, Canada

**Keywords:** COVID-19, SARS-CoV-2, critical, Long COVID, Africa, low-income country, immune activation, inflammation, proteomics

## Abstract

Little is known about the acute and long-term sequelae of COVID-19 and its pathophysiology in African patients, who are known to have a distinct immunological profile compared to Caucasian populations. Here, we established proteomic signatures to define severe outcomes of acute COVID-19 and determined whether proteome signatures during the first week of acute illness predict the risk of post-acute sequelae of COVID-19 (Long COVID) in a low-income country (LIC) setting. Using the Olink inflammatory panel, we measured the expression of 92 proteins in the plasma of COVID-19 patients (n=55) and non-COVID-19 individuals (n=23). We identified distinct inflammatory proteomic signatures in acute severe COVID-19 individuals (n=22) compared to asymptomatic or mild/moderate COVID-19 cases (n=33), and non-COVID-19 controls. Levels of SLAMF1, CCL25, IL2RB, IL10RA, IL15RA, IL18 and CST5 were significantly upregulated in patients with critical COVID-19 illness compared to individuals negative for COVID-19. The cohort was followed for an average of 20 months, and 23 individuals developed Long COVID, based on the WHO’s case definition, while 32 COVID-19 patients recovered fully. Whereas upregulated levels of SLAMF1, TNF, TSLP, IL15RA, IL18, ADA, CXCL9, CXCL10, IL17C, and NT3 at the acute phase of the illness were associated with increased Long COVID risk, upregulated expression of TRANCE was associated with a reduced risk of developing Long COVID. Proteomic expression levels of SLAMF1, IL15RA, and IL18 associated with critical illness during the acute phase of COVID-19 also predicted Long COVID risk. Unravelling the pathophysiology of severe acute COVID-19 and Long COVID before its advent may contribute to designing novel interventions for diagnosing, treating, and monitoring of SARS-CoV-2 infection and its associated acute and long-term consequences.

## Introduction

Acute COVID-19 is characterized by protean clinical manifestations, including asymptomatic, mild/moderate, severe, or critical conditions. Moreover, a multitude of complex symptoms persist in SARS-CoV-2 infected individuals post-acute phase, affecting the cardiovascular, respiratory, gastrointestinal, genitourinary, hematologic, musculoskeletal, central nervous, and other systems, collectively known as post-acute sequelae of COVID-19 or Long COVID (1,2). The WHO defines Long COVID as an ongoing, relapsing, or new symptom or condition present three or more months from the onset of COVID-19 with symptoms that last for at least two months and cannot be explained by an alternative cause (3). However, other organizations use different definitions for Long COVID indicating the elusive characteristics of the condition (4,5). Systematic reviews demonstrated that the burden of Long COVID ranges between 45% and 62% depending on the case definition, the study design, and the region where the study was conducted (6–8). Most of these data have been reported from high-income countries (HICs), and a significant knowledge gap exists in low-income country (LIC) settings (9–10).

Though its exact cause remains unknown, age, sex, socio-economic conditions, ancestry, co-morbidities, severity during acute illness, reinfection, SARS-CoV-2 genotype, and vaccination appear to be associated with the risk of developing Long COVID (1,2,11–16). Persistence of replication-competent viruses, or viral components, reactivation of latent virus, autoimmunity, microbiota dysbiosis, and chronic inflammation have been proposed as the mechanism(s) leading to multiple organ damage in patients with Long COVID (2). Several biomarkers have been investigated to assess the risk of severe illness during acute COVID-19 or Long COVID in HICs (17–38). However, many of these biomarkers have not been validated and are not yet commonly used in clinical practice. Additionally, there is limited knowledge of the overall pathophysiology and molecular mechanisms underlying the acute and long-term sequelae of COVID-19 in LICs. In particular, the distinct immunological background, high burden of co-infections due to HIV-1, malaria, tuberculosis, helminths, and the diverse sociodemographic factors may impact the biological profile following infection with SARS-CoV-2 infection in African populations (39–42). Indeed, a recent report demonstrated distinct COVID-19 immune signatures associated with COVID-19 severity in Ugandan patients co-infected with HIV-1 (21). Given that Long COVID is a heterogeneous disease with complex symptoms, it is imperative to unravel the role of biomarkers that can predic the development of severe acute COVID-19 and Long COVID in the context of Africa.

In this study, we performed a longitudinal analysis of patients in Ethiopia with COVID-19. Specifically, we compared proteomic profiles between COVID-19 patients and COVID-19-negative individuals. In addition, we assessed plasma proteomic signals associated with severe outcomes following acute COVID-19 and determined whether unique proteins that appear early during the onset of acute COVID-19 illness predict Long COVID risk. Our findings show that differential expression of proteomes represents immune dysregulation in COVID-19 individuals who develop critical illness as well as those who developed Long COVID. Understanding the pathophysiology of acute COVID-19 illness, and Long COVID before its advent may contribute to designing novel interventions related to diagnosing, treating, and monitoring acute COVID-19, Long COVID, and other chronic post-viral syndromes.

## Results

### Study participants and characteristics

Study participants were recruited from an ongoing prospective observational cohort study in Ethiopia (43). A cohort of 55 patients with confirmed COVID-19 were recruited for this study. In addition, another 23 randomly selected COVID-19-negative individuals examined for other respiratory illnesses were enrolled as controls (Figure 1). There were no significant differences in sociodemographic or clinical factors, SARS-CoV-2 vaccine uptake, hospital admission status or medication taken among the two groups (Table 1, Figure 2A). However, only COVID-19 patients were admitted to the intensive care unit (ICU).

**Fig. 1.**
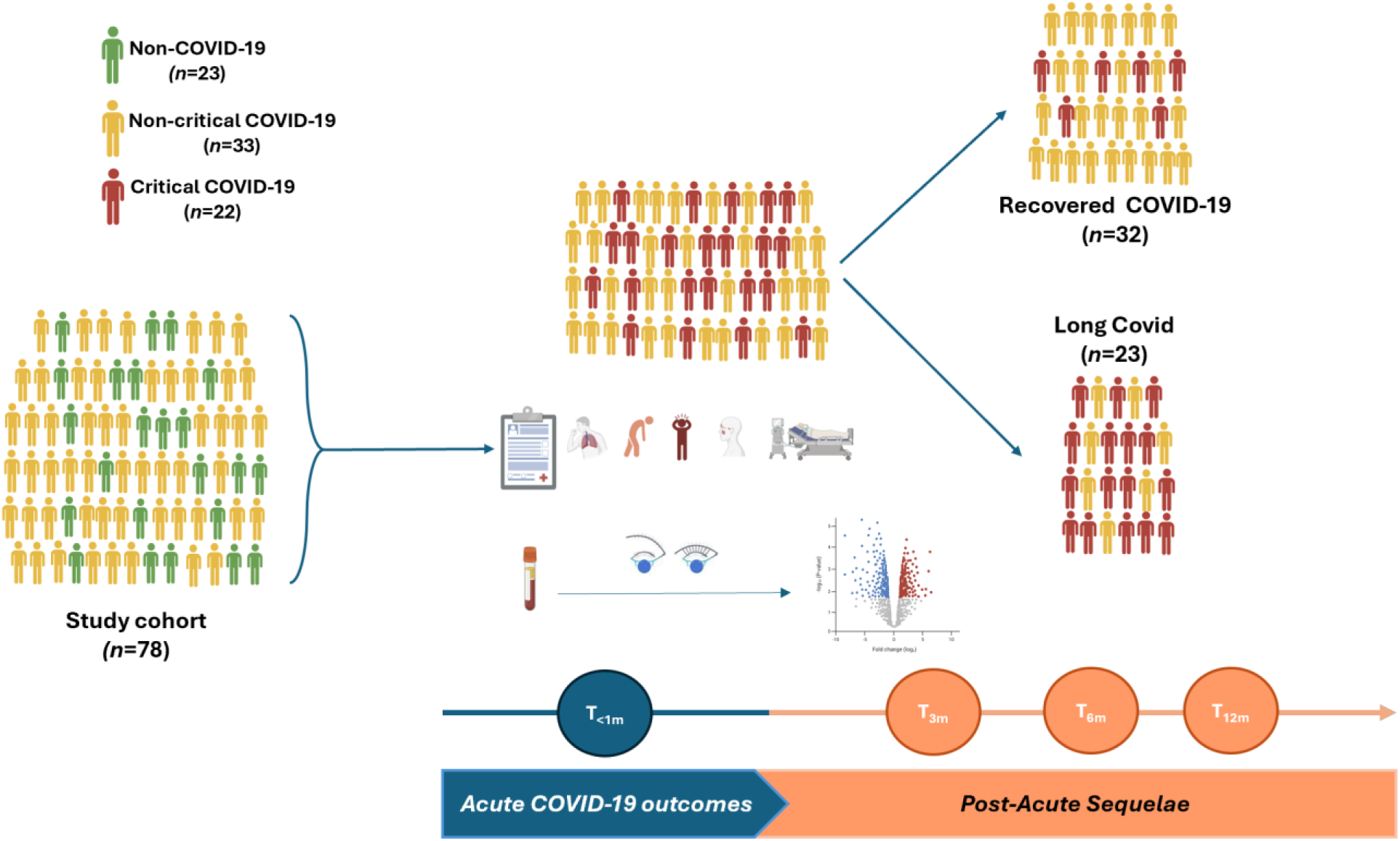
Study design and data analysis.

**Fig. 2.**
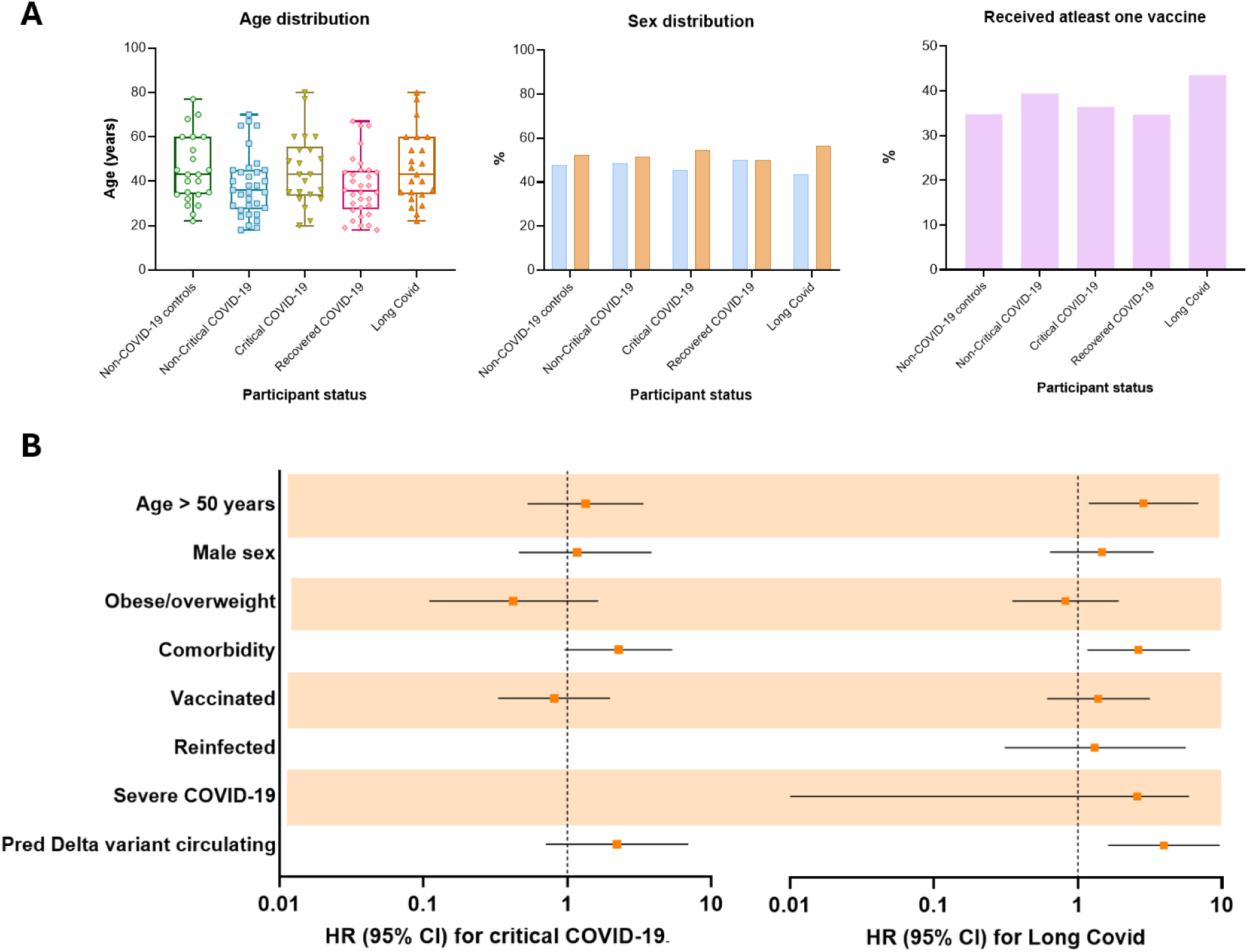
Clinical characteristics of study groups at baseline. **(A)** Age-, sex-distribution, and SARS-CoV-2 vaccination coverage of the different group study participants. (B) Adjusted hazard risk (HR) for critical illness during the acute phase of COVID-19, or Long COVID.

**Table 1.** Baseline Sociodemographic and Clinical Characteristics of Study Participants.

| Characteristic | Non-COVID-19 controls<br>(n = 23) | COVID-19 positive |  |  |  |  |
| --- | --- | --- | --- | --- | --- | --- |
|  |  | All COVID-19<br>(n =55) | Acute illness outcome |  | Long-term outcome |  |
|  |  |  | Non-critical<br>(n=33) | Critical<br>(n=22) | Recovered<br>COVID-19<br>(N=32) | Long COVID<br>(n=23) |
| Sex (Female) <sup>§</sup> | 11 (47.8) | 37 (47.4) | 16 (48.5) | 10 (45.5) | 16 (50.0) | 10 (43.5) |
| Age [median, y (IQR)] | 43 (34-60) | 38 (29-49) | 36 (28-45) | 43 (34-54) | 35.5 (28-45) | 43 (34-60) |
| Age group |  |  |  |  |  |  |
| ≥50 | 7 (30.4) | 12 (21.8) | 5 (15.2) | 7 (31.8) | 4 (12.5) | 8 (34.8)* |
| BMI category<br>(overweight and obese) | 9 (39.1) | 21 (38.2) | 14 (42.4) | 7 (31.8) | 13 (40.6) | 8 (34.8) |
| Comorbidity<br>(at least 1 pre-existing condition) | 7 (30.4) | 17 (30.9) | 4 (12.1) | 13 (59.1)**** | 6 (18.8) | 11 (47.8)* |
| Hospital admission | 9 (39.1) | 22 (40.0) | 0 (0.0) | 22 (100.0)*** | 6 (18.8) | 16 (69.6)**** |
| Intensive Care Unit admission | 0 (0.0) | 7 (12.7)**** | 0 (0.0) | 7 (31.8)*** | 4 (12.5) | 3 (13.0) |
| Vaccination (at least 1 dose) | 8 (34.8) | 21 (38.2) | 13 (39.4) | 8 (36.4) | 11 (34.4) | 10 (43.5) |
| On medications |  |  |  |  |  |  |
| Anticoagulants | 7 (30.4) | 18 (32.7) | 0 (0.0) | 18 (81.8)*** | 5 (15.6) | 13 (56.5)** |
| Antibiotics | 8 (34.8) | 24 (43.6) | 6 (18.2) | 18 (81.8)*** | 12 (37.5) | 12 (52.2) |
| Immunosuppressants | 7 (30.4) | 14 (25.5) | 0 (0.0) | 14 (63.6)**** | 3 (9.4) | 11 (47.8)** |
| Oxygen supplemental therapy | 9 (39.1) | 22 (40.0) | 0 (0.0) | 22 (100.0)*** | 6 (18.8) | 16 (69.6)*** |
| Predominant variant in pandemic wave |  |  |  |  |  |  |
| Delta | N/A | 22 (40.0) | 4 (12.1) | 18 (81.8)**** | 9 (28.1) | 13 (56.5)* |
| Omicron | N/A | 33 (60.0) | 29 (87.9) | 4 (18.2) | 23 (71.9) | 10 (43.5) |
| SARS-CoV-2 reinfection | N/A | 5 (9.1) | 4 (12.1) | 1 (4.6) | 3 (9.4) | 2 (8.7) |

<sup>§</sup>Data are participants, No. (%) unless otherwise described.
\*\*\*p<0.001; \*\*\*\*p<0.0001 (Pearson's Chi2 or Fisher's exact test, where appropriate) when non-COVID-19 compared to COVID-19, or non-critical compared to critical COVID-19 patients, or recovered compared to Long COVID.

Of the 55 individuals with COVID-19, 33 had mild or moderate (non-critical) disease and 22 were severe, requiring hospital admission only or with intensive care (critical). Fatigue/malaise (54.6%), arthralgia (52.7%), myalgia (50.9%), fever (50.9%), loss of smell (49.1), sore throat (47.3%), loss of taste (45.5%), headache (45.5%), anorexia (45.5%), cough (40.0%), and shortness of breath (30.9%) were the most frequent symptoms experienced by COVID-19 patients during the acute phase of COVID-19 illness (Table 1 supplementary). Critically ill COVID-19 cases were older than non-critical patients, had significantly more frequent symptoms, more comorbid conditions, as well as more frequent hospital and intensive-care unit (ICU) admissions. Anticoagulants, antibiotics, anti-inflammatory drugs, and oxygen supplementation were more frequently administered to critically ill COVID-19 patients than non-critical COVID-19 counterparts. SARS-CoV-2 Delta and Omicron were the two most predominantly circulating variants during the pandemic wave in Ethiopia when the study was undertaken (Supplementary Figure 1) (44–46). As expected, the predominant circulating SARS-CoV-2 variant (i.e. Delta) at the time of enrollment was significantly associated with critical acute COVID-19 illness (Table 1).

Participants were followed for an average of 20 (IQR: 9-21) months. Overall, 41.8% (23 out of 55) of COVID-19 patients reported experiencing at least one persistent symptom, according to the WHO’s case definition for Long COVID (3). The most commonly reported symptoms among these patients were fatigue (47.8%), cough (47.8%), myalgia (47.8%), insomnia (30.4%), foggy brain (21.7%) and shortness of breath (21.7%). Notably, age > 50 years (HR=2.85, 95% CI: 1.19-6.86), having at least one comorbid condition (HR=2.63, 95% CI: 1.16-5.99), and being infected during the predominant circulating Delta SARS-CoV-2 variant phase of the pandemic wave (HR=2.58, 95% CI: 1.13-5.91) were associated with a significantly increased hazard of Long COVID (Figure 2B).

### COVID-19 patients exhibited increased levels of inflammatory proteins

Overall, 14,076 protein levels in 153 samples (78 at baseline, 44 at 6-month and 31 at 12-month follow-up) derived from 78 individuals were measured using the Olink targeted 96 inflammation proximal extension assay (PEA) platform. Initially, we compared the proteomic signatures of COVID-19 patients during the acute phase of COVID-19 illness (n=55) with the COVID-19-negative non-COVID-19 controls (n=23). Figure 3A shows the clustering of inflammatory proteome profile in individuals with COVID-19 compared to non-COVID-19 controls. Thirty-one proteins were significantly increased in COVID-19 patients compared to non-COVID-19 controls after adjustment for multiple testing (Figure 3B and 3C). The top ten significantly increased proteins were TNF, IL7, VEGFA, CCL20, CSF1, CXCL6, MCP4, CD40, FGF21, and CXCL11 (Figure 3C).

**Fig. 3.**
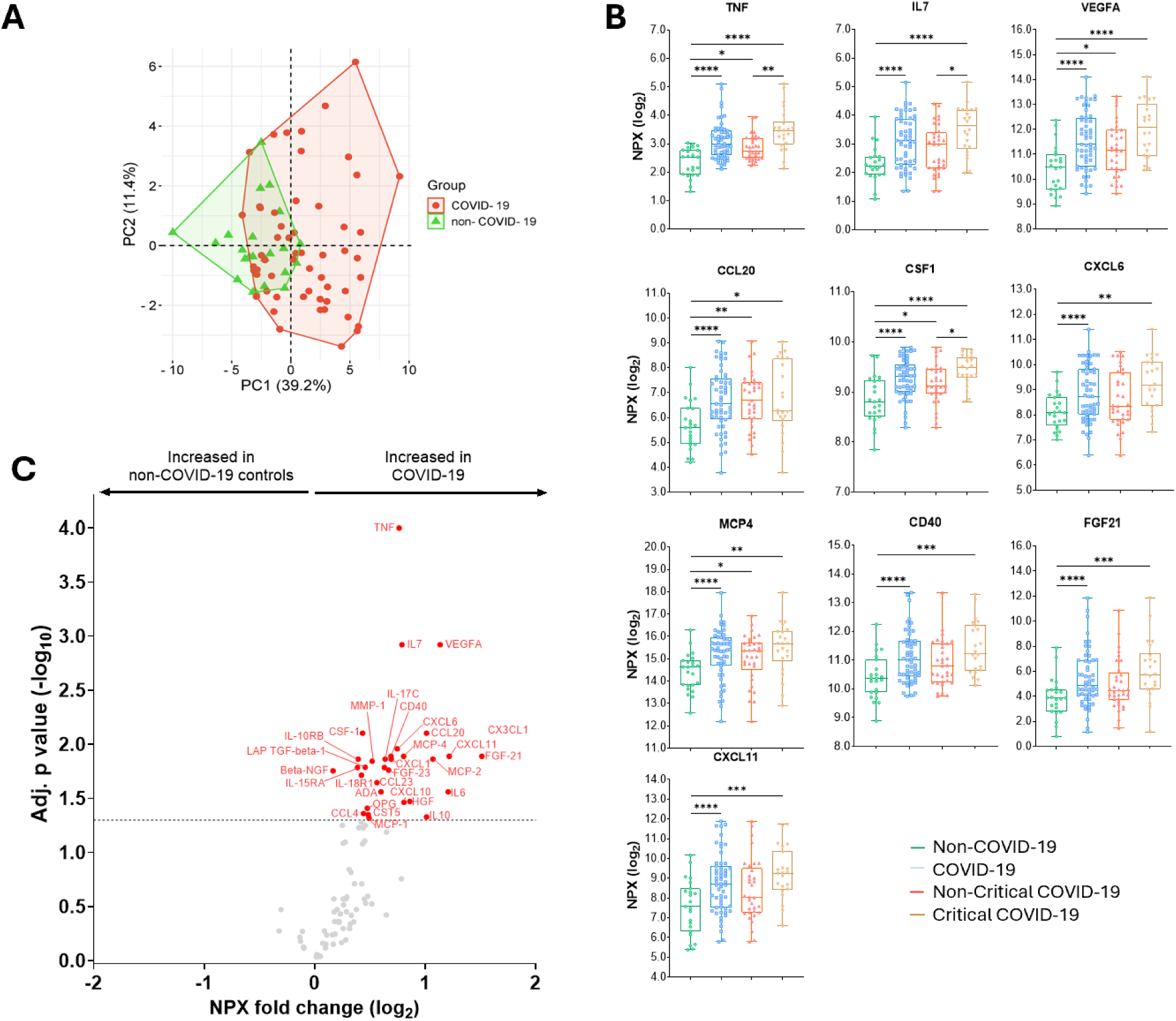
Differential expression of inflammatory proteome in COVID-19 patients compared to non-COVID-19 controls. **(A)** A principal component analysis between COVID-19 and non-COVID-19 individuals. **(B)** Median (IQR) protein expressions [expressed as log_2_-transformed Normalized Protein eXpression (NPX) values in non-COVID-19 controls, COVID-19, non-critical COVID-19, or critical COVID-19 patients. P values: *<0.05, **0.01, ***<0.001, and ****<0.0001 were calculated using Kruskal Wallis with Dun’s correction for multiple tests. **(C)** A volcano plot showing differential expression analysis comparing proteomics from COVID-19 patients with non-COVID-19 controls. Red and blue dots represent significantly (adjusted for multiple testing by Benjamini-Hochberg) up- and down-regulated proteomics, respectively. Gray dots indicate nonsignificant.

### COVID-19 patients with critical illness exhibit a distinct plasma proteomic signature

Within the COVID-19 patient group, we then determined the proteomic signatures in those with critical outcomes (n=22) and compared them with those who did not develop critical COVID-19 (n=33), or non-COVID-19 controls (Figure 3B, 4A-D). We identified that only three proteins, namely TNF, SLAMF1, and CDCP1, were differentially expressed in critically ill COVID-19 patients compared to those with non-critical illness presentation (Figure 4A). However, when critical COVID-19 patients were compared to non-COVID-19 controls, there was significant clustering between the two groups (Figure 4B), and the number of differentially expressed proteins increased significantly to 31 (Figure 4C and 4D). Seven proteins that were highly expressed and associated with an increased hazard of critical COVID-19 outcome were SLAMF1, CCL25, IL2RB, IL10RA, IL15RA, IL18 and CST5 (Figure 4E). Additionally, the predictive power assessed by the AUC curve showed that IL15RA (AUC=0.842), SLAMF1 (AUC=0.775) and IL18 (AUC=0.775) as the top three proteins predicting critical COVID-19 illness (Figure 4F). The combined AUC score of these three proteins is 0.836 (Supplementary Figure 3A). Gene Ontology (GO) analysis revealed significant enrichment in phagocytotic pathway, and protein-protein interaction pathways showed significant cytokine-cytokine, cytokine-cytokine receptor, and cytokine-chemokine, cytokine-chemokine receptor, chemokine-chemokine receptor interactions when critical and non-critical COVID-19 patients were compared (Figure 4G).

**Fig. 4.**
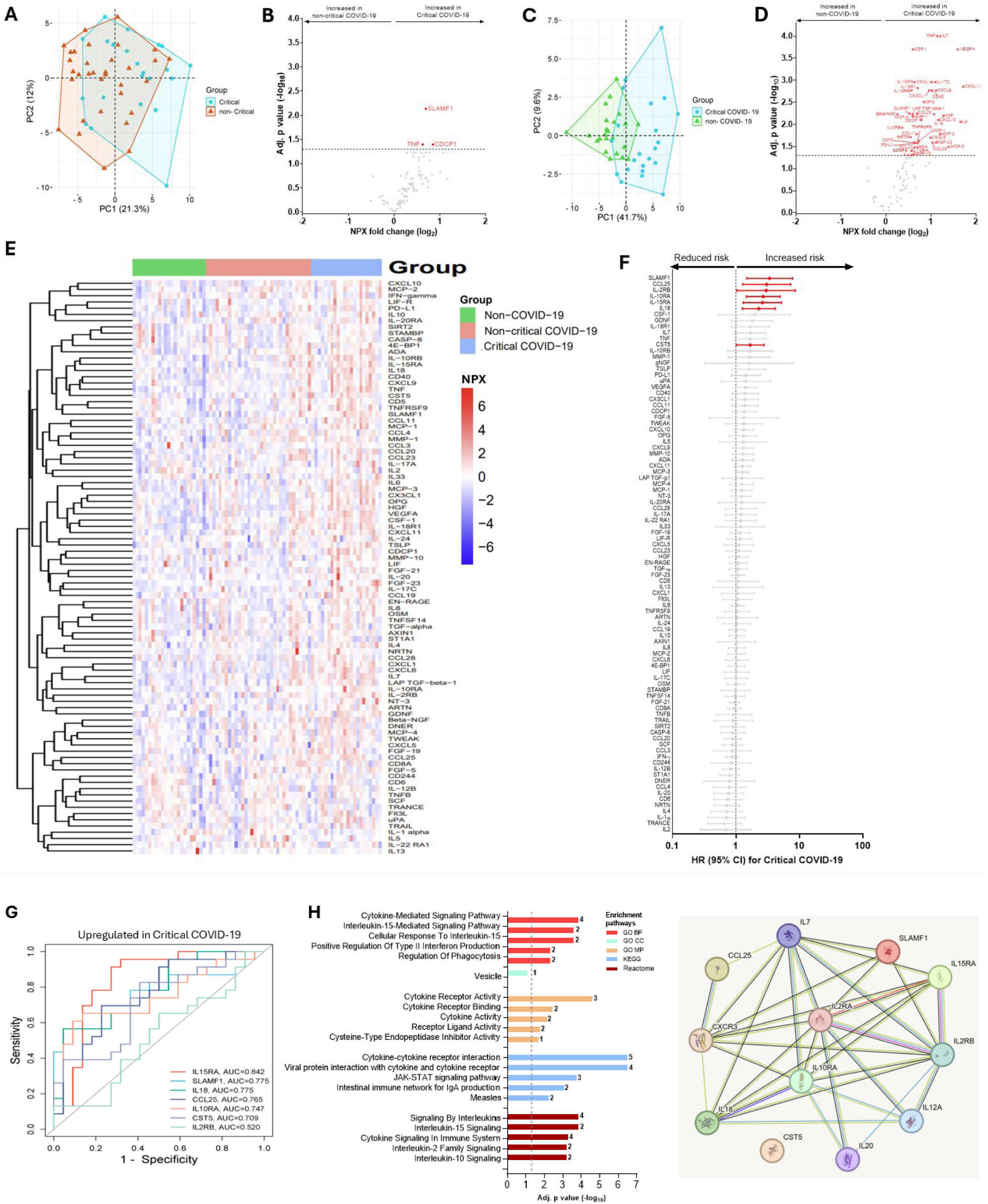
Differential expression of inflammatory proteome in critical COVID-19 patients compared to non-critical COVID-19 patients or non-COVID-19 controls. Predicting power of differentially expressed proteins, gene enrichment and protein-protein interactions. A principal component analysis between critical COVID-19 and non-critical illness **(A),** or between critical COVID-19 and non-COVID-19 individuals **(C)**. A volcano plot showing differential expression analysis comparing proteomic expression levels of critical COVID-19 patients with non-critical COVID-19 patients **(B)**, or non-COVID-19 controls **(D)**. Red dots represent significantly (adjusted for multiple testing by Benjamini-Hochberg) upregulated proteins. Gray dots are insignificant. **(E)** Heat maps showing differential expression of proteins in individuals without COVID-19, non-critical COVID-19, or critically ill COVID-19 **(F)** Adjusted cox-proportional hazard risk (HR) for critical illness. HR [(95% confidence intervals (CI)] in red colour signifies increased risk of critically ill COVID-19 patients per unit change in log_2_-transformed NPX values of each protein. Gray colours are statistically not significant values. **(G)** Receiver operating characteristic (ROC) curve analysis of the upregulated proteins associated with Critical COVID-19 illness. **(H)** Gene enrichment and protein-protein interaction pathways of significantly expressed proteomes between critical and non-critical COVID-19 patients.

### Differential plasma proteomic expression during acute COVID-19 illness predicts the risk of Long COVID

Here, we compared proteomic signatures in the study cohort with long-term outcomes – namely Long COVID. Proteomic expression levels in recovered COVID-19 patients segregated clearly from those who eventually developed Long COVID (Figure 5A). Overall, COVID-19 patients who developed subsequent Long COVID exhibited 44 differentially expressed proteins compared to those who recovered (Figure 5B, 5D). Of these, ten proteins, namely SLAMF1, TNF, TSLP, IL15RA, IL18, ADA, CXCL10, IL17C, NT3, and CXCL9 were significantly increased (Figure 5C), and associated with an increased hazard of developing Long COVID after controlling for age, comorbidity and the Delta variant (Figure 5E). Notably, the expression level of the protein TRANCE was reduced in patients who developed Long COVID compared to recovered COVID-19 patients and reduced the Long COVID risk (Figure 5C-5E). After adjusting for age, comorbidity and SARS-CoV-2 variant, the hazard of Long COVID was 1.58 up to 4.12-fold in COVID-19 patients with upregulated baseline protein expression levels of SLAMF1, TNF, TSLP, IL15RA, IL18, ADA, CXCL10, IL17C, NT3, and CXCL9 (Figure 5E). On the contrary, it was only 0.50-fold in patients with a higher baseline TRANCE expression levels (Figure 5E). Additionally, the predictive power assessed by the AUC curve showed that TNF (AUC=0.973), SLAMF1 (AUC=0.894) and IL18 (AUC=0.857) as the top three proteins predicting increased Long COVID, and TRANCE (AUC=0.700) predicted reduced Long COVID risk (Figure 5F). The combined predictive AUC score of these three proteins in predicting Long COVID is 0.857 (Supplementary Figure 3B). Gene Ontology (GO) analysis revealed significant enrichment in cytokine-, chemokine-, IL17-, TLR-signaling pathways, and protein-protein interaction pathways showed significant cytokine-cytokine, cytokine-cytokine receptor, and cytokine-chemokine, and chemokine-chemokine interactions when recovered COVID-19 and Long COVID patients were compared (Figure 5G).

**Fig. 5.**
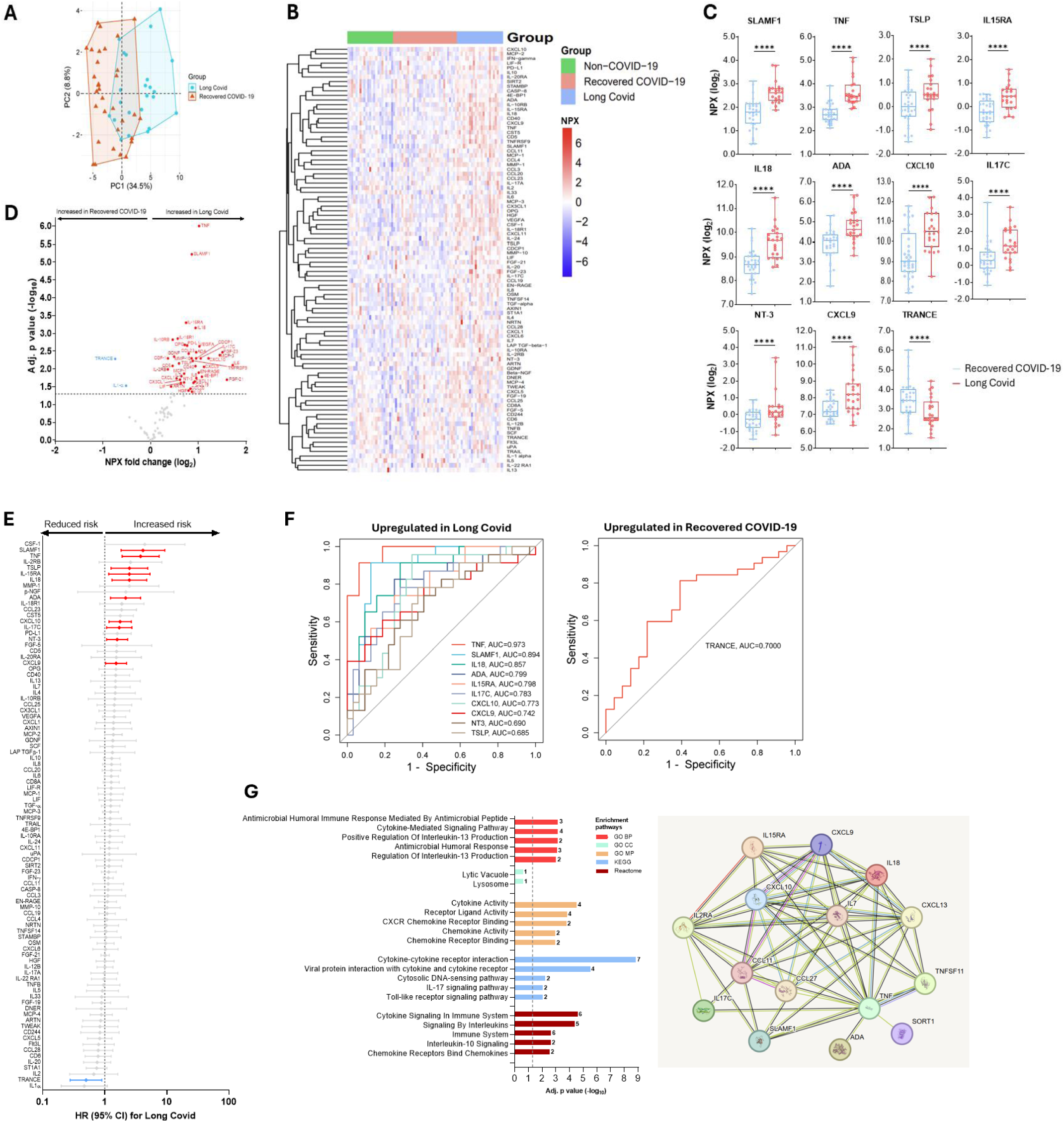
Differential proteomic expression in individuals with COVID-19 who developed Long COVID compared to recovered COVID-19 patients, and predicting power of differentially expressed proteins, gene enrichment and protein-protein interactions of significantly differed proteins in Long COVID. **(A)** A principal component analysis between recovered COVID-19 and those who developed Long COVID. **(B)** Heat maps showing differential expression of proteins in non-COVID-19, recovered COVID-19, or Long COVID. **(C)** Median (IQR) protein expressions in individuals with COVID-19 who developed Long COVID compared to recovered COVID-19 patients. P values, ****<0.0001 were calculated using Kruskal Wallis with Dun’s correction for multiple tests. **(D)** A volcano plot showing differential expression analysis comparing proteomics in those who developed Long COVID with those who recovered. Red and blue dots represent significantly (adjusted for multiple testing by Benjamini-Hochberg) up- and down-regulated proteins, respectively. Gray dots indicate nonsignificant. **(E)** Adjusted cox-proportional hazard risk (HR) for Long COVID. HR (95% CI) in red signifies an increased risk and blue signifies a reduced risk of Long COVID per unit change in log_2_-transformed NPX values of each proteome. Gray colours are not significant. **(F)** Receiver operating characteristic (ROC) curve analysis of the upregulated or downregulated proteins associated with Long COVID risk. **(G)** Gene enrichment and protein-protein interaction pathways of significantly expressed proteomes between critical and non-critical COVID-19 patients.

### Differential proteomic signatures persist throughout post-acute sequelae of COVID-19

During the first year period following enrollment, plasma samples were obtained in a subset of recovered COVID-19 patients and those who developed Long COVID at an average of 6- and 12-months. Trends in the profile of proteomic expression were distinct between the two groups (Figure 6B). Although we observed a tendency towards reduction, the majority of the proteins associated with the increased risk of Long COVID remained unchanged during the follow-up period, except TNF and IL18 which showed significant reductions. On the contrary, IL17C and NT3 levels increased significantly among recovered COVID-19 patients. CXCL10 was the only protein that exhibited a significant reduction in both recovered and Long COVID patients. There was a tendency towards increases in TRANCE levels in both recovered and Long COVID cases, the difference was not significant.

**Fig. 6.**
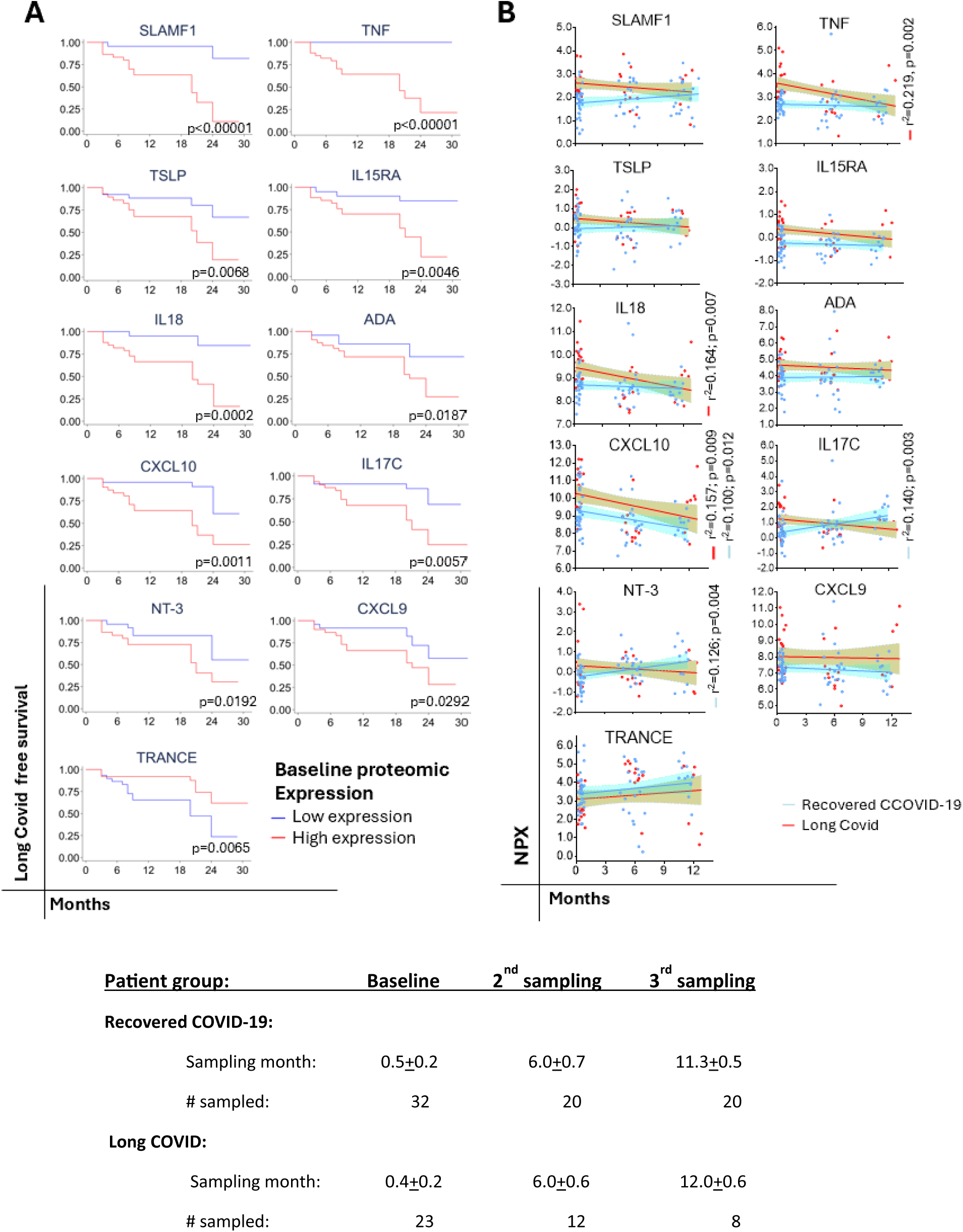
Predicting of progression to Long COVID and trajectories of proteomic expression levels overtime. **(A)** Kaplan-Meier curves showing the clinical progression of Long COVID. Statistical differences between high (≥ median NPX) or low (< median NPX) proteomic expression levels were estimated by rank test. **(B)** Simple regression analysis of the trajectories of the significantly expressed proteomic levels in recovered COVID-19 versus Long COVID.

## Discussion

In this study, we demonstrated that circulating proteome is broadly dysregulated in COVID-19 patients and is associated with the risk of post-COVID-19 sequelae. To the best of our knowledge, the current study is the first to characterize the proteomic profiles of acute and long-term COVID-19 outcomes in the context of Sub-Saharan Africa.

Acute COVID-19 is characterized by immune activation and inflammation (47). Our data is consistent with other studies that find that COVID-19 patients exhibit increased levels of 31 unique proteins compared to non-COVID-19 controls (17–27). This includes proteins such as TNF, CD40, CSF, IL6, IL10, IL18RA, CCL23, CXCL10, CXCL11, MCP-2, and FGF21, which has been reported previously (17–27). Dysregulation of these cytokines and chemokines has been related to excessive inflammation and tissue damage in COVID-19 patients, particularly among COVID-19 patients with severe clinical outcomes (47). Here, we demonstrated that the unique signature featuring SLAMF1, CCL25, IL2-RB, IL10RA, IL15RA, IL18, and CST significantly increased the risk of critical illness in COVID-19 patients. Increased levels of SLAMF1 and IL18 in this patient group were similar to those reported previously (17–21). However, other investigators reported that CCL25 was lower in ICU-admitted patients than in non-ICU cases, and increased CCL25 was observed with clinical improvement (22). We were not able to confirm the findings in our setting; however, a major difference is that our study is derived from a setting in a LIC. Overall, these inflammatory response-related cytokines and chemokine ligands play a pivotal role in the pathogenesis of tissue injury associated with severe COVID-19.

High expression of the proteins SLAMF1, TNF, TSLP, IL15RA, IL17C, IL18, ADA, CXCL9, CXCL10 and NT3 during the acute phase of the illness was associated with an increased hazard of Long COVID in our cohort. Similar to our data, previous findings also showed increased levels of TNF, IL18, CXCL9, and CXCL10 (20,30,31). Notably, baseline protein expression levels of SLAMF1, IL15RA and IL18 associated with the critical illness during the acute phase of COVID-19 were also able to predict Long COVID risk. These biomarkers may play a pivotal role in the earlier identification of individuals who will succumb to severe disease during the acute phase of the illness as well as long-term sequelae. TSLP is a cytokine involved in the context of inflammation and allergy. Recent studies have shown that TSLP levels correlated with the duration of hospitalization in COVID-19 patients (48). This suggests that TSLP might play a role in the prolonged inflammatory responses seen in Long COVID. Most of the proteins associated with the increased Long COVID risk remained unchanged during the follow-up period indicating a sustained immune activation and inflammation. However, three proteins, namely TNF, IL18, and CXCL10, exhibited significant reductions, and the repeated measurement of these biomarkers may be relevant in monitoring treatment outcomes. Whereas studies concur with our result showing continuous reductions at 6 months of follow-up (30), others showed persistent elevations up to 18-24 months (20). In addition, persistent and sustained level of CXCL9 was observed in the current study as reported by others (20). Two proteins, namely IL17C and NT3 exhibited significant increased expression levels among recovered COVID-19 patients. IL17C is a member of the IL17 family that plays a role in the immune response and enhances inflammatory responses by inducing the release of cytokines, such as IL1β, TNFα and IL6 (49). It might play a role in the persistent activation of the immune system contributing to the symptoms observed in Long COVID. NT3, a cytokine primarily known for its role in the nervous system (50), could be relevant in Long COVID through its role in immune regulation and tissue repair. By influencing the activity and survival of immune cells, NT3 might help modulate the persistence of immune activation and inflammation seen in Long COVID.

TRANCE, also known as RANKL, or TNFSF11, is known to be involved in the regulation of T cells and dendritic cells (51), which are key players in the immune response against SARS-CoV-2 (47). In this study, a high baseline level of TRANCE was associated with a reduced risk of developing Long COVID. Notably, reduced expression of TRANCE has been associated with severe COVID-19, including admission to the ICU (19,26), and low levels of TRANCE in the CSF predicted Long COVID in patients followed for 13 months (52). Given that Long COVID has been linked to persistent viral replication (53–56), it is tempting to propose that this biomarker plays a role in modulating the anti-SARS-CoV-2 immune response.

The differences in the findings between our data and other studies may be attributed to differences in the case definitions for COVID-19 severity, Long COVID, follow-up duration, study design, and differences in patient population characteristics. We and others have previously demonstrated that acute COVID-19 illness in Africa is, in general, less severe than in HICs despite the intense transmission rate in LICs (11,57,58). Notably, an estimated 25-42% of COVID-19 individuals in HICs end up being hospitalized or admitted into the intensive care unit (ICU), particularly during the earlier phases of the pandemic due to the ancestral variant (59–61). On the contrary, in the setting of LICs, only <5% of all COVID-19 patients develop severe COVID-19 (11,57). This has been ascribed to pre-existing cross-immunity, immuno-modulation, or trained immunity (39–41). Likewise, we hypothesize that a similar mechanism operates in the pathogenesis of Long COVID showing a low burden of Long COVID in certain parasite-endemic areas in Africa (62).

A strength of our study includes the interrogation of proteomics related to the pathophysiology of acute and long-term sequelae of SARS-CoV-2 infection in an LIC setting. Additionally, the plasma sampling time and the assessment of the baseline proteomic levels for predicting Long COVID risk. Several studies have included samples that were analyzed ranging between day <1 and >24 months (63–65). Adjustment for confounders which included age, comorbidity and SARS-CoV-2 variant associated with increased risk of Long COVID in our cohort is another strength of this study. Finally, albeit in a small number of patients, we also included the assessment of protein level trajectories over time that revealed some unique features that may help in Long COVID treatment outcomes. Nonetheless, our study has several limitations. Similar to several other studies, our cohort also suffers from analysis based on a relatively small sample size (63–65). Second, we did not have SARS-CoV-2 genotype data although we have attempted to relate our analysis to the predominant variants circulating in the country during the enrollment period of our cohort participants (44–46). Finally, we determined 92 proteins focusing on inflammatory panels and we may have failed to detect all relevant proteomic biomarkers.

In conclusion, our data revealed that unique proteomic signatures are associated with severe COVID-19 and Long COVID in the African context. Dysregulation of proteomic patways involved in cellular degranulation and proteolysis was significantly expressed among COVID-19 patients versus non-COVID-19 controls. Additionally, significantly increased unique proteomic expression in critically ill COVID-19 patients compared to non-COVID-19 controls or recovered COVID-19 patients compared to those who developed Long COVID exhibited inflammatory cytokines, chemoattractant for neutrophils, T cells, NK cells, monocytes, and endothelial cells. The findings should serve as baseline data informing future -omics studies in LMIC settings involving a larger sample size and more diverse additional proteomic panels related to cardiovascular, neurological, and metabolic dysregulations associated with post-COVID-19 sequelae (32–36). Additionally, this study provides critical insights into the pathophysiology of acute and long-term consequences of COVID-19, informing strategies and treatment approaches (66).

## Methods

### Patient recruitment and follow-up

This study (Clinicaltrials.gov: NCT04584424) is a prospective observational cohort study being undertaken in Ethiopia and the study protocol has been described in detail previously (43). The study was reviewed and approved by the Health Research Ethics Review Committee of the Ethiopian Public Health Institute in Ethiopia (EPHI-IRB-282-2020) and the Hamilton Integrated Research Ethics Board (HiREB:16956). Written informed consent was obtained by all participants, or their guardians, for participation in the study.

Adults 18 years and older presenting with respiratory infections attending hospital-based settings between August 2021 and February 2022 were recruited and screened using real-time polymerase chain reaction (RT-PCR) and/or antigen tests, or anti-nucleocapsid antibody tests. Individuals with confirmed SARS-CoV-2 infection (i.e., positive in any of the tests mentioned above) were considered COVID-19 cases. Those with a negative SARS-CoV-2 PCR, or antigen, or no documented evidence of COVID-19 clinically were considered as COVID-19-negative non-COVID-19 controls. The controls were not tested for other pathogens although the patients had “influenza-like illness” at enrollment. Individuals without evidence of COVID-19 at enrollment who became infected during follow-up were excluded. In addition, individuals with immunosuppression, including HIV-1 infection were excluded. Sociodemographic, clinical, and laboratory data were collected using standardized Case Record Forms (CRFs) adapted from the International Severe Acute Respiratory and Emerging Infection Consortium’s (ISARIC) CRFs for emerging severe acute respiratory infections (67). Data was entered using the REDcap software package. The baseline patient’s severity status was classified following the WHO criteria as asymptomatic, mild/moderate, severe (with dyspnea, respiratory rate ≥ 30 breaths per minute, O2 saturation ≤ 93%, lung infiltrates ≥ 50% of the lung fields within 24-48 hours), and critical (with respiratory failure, septic shock, and/or multiple organ failure) (68). Cohort follow-up was conducted for the survey of persistent symptoms occurring at any time from the onset of COVID-19 included in the WHO case definition and defined as the presence of at least one persistent symptom of > 2 months duration occurring 12 weeks from the onset of acute COVID-19 illness (3).

### Sample collection and processing

Peripheral blood was collected using EDTA vacutainers within 1 to 21 days of SARS-CoV-2 diagnosis during acute illness, and follow-up samples were collected after 180 (±30), and 365 (±30) days post-symptom onset. Plasma was stored frozen at −80°C until analysis and transported to McMaster University for Olink proteomic analysis.

### Proteomic analysis

The Olink targeted 96 inflammation proximal extension assay (PEA) platform (Uppsala, Sweden) was used as per the manufacturer’s guidelines (69). The Olink Target 96 Inflammation panel offers a broad selection of proteins associated with inflammatory and immune response processes. In brief, the PEA platform uses antibody pairs linked to unique DNA oligonucleotides, that bind to target proteins. The binding of the antibodies to their target proteins brings the DNA oligonucleotides into closer proximity and results in the hybridization and formation of a new DNA sequence. The DNA sequence is then amplified and quantified using real-time PCR. Normalized Protein eXpression (NPX), an arbitrary unit normalized into the log_2_ scale, is used to define the protein expression level. Measurements that failed the internal quality control with a warning were excluded from the dataset.

### Statistical analysis

Baseline characteristics with continuous variables were summarized as median [interquartile ranges (IQR)], and categorical variables as frequencies (percentages). Continuous variables were compared by Mann-Whitney U or Kruskal-Wallis tests, and categorical variables using χ² test or Fisher’s exact test. Cox proportional hazard (HR) was used to ascertain the association between explanatory variants (including age, sex, body-mass index, comorbidity, vaccination, reinfection, SARS-CoV-2 variant) and outcomes of interest, namely critical COVID-19 during acute illness and development of Long COVID. Multivariate HR was estimated by including all significant values (age above 50 years, comorbidity, and infection during the predominant circulating Delta variant) in univariate analysis.

Initial unsupervised clustering of groups was performed by principal component analysis (PCA) and heat maps. The differential fold-change (log_2_) expression of proteomics was estimated between the different groups. Multiple testing corrections were applied according to Benjamini and Hochberg’s procedure, and data were visualized using violin plots. The association between the expression of each protein and outcome (i.e. critical illness or Long COVID) was then compared using cox-proportional hazard. Cox proportional regression models were conducted to estimate the association of each biomarker expression level [as the dependent variable, categorized into high (≥ median NPX) or low (< median NPX) expression levels] with incident Long COVID followed by adjustments for confounding covariates (70). Kaplan-Meier curves were created to visualize the effect of protein expression levels on the risk of developing Long COVID. Statistical differences between groups were estimated by rank test. Gene Ontology (GO) terms and STRING was used for protein-protein interaction pathways analysis (70). In addition, Receiver operating curves (ROCs) was used to determine which protein significantly predicted symptoms. p-values < 0.05 were considered significant. R studio, GraphPad Prism, and STATA software were used for statistical analysis.

## Acknowledgments

This work received grants from the Ethiopian Federal Ministry of Health, Addis Ababa, Ethiopia, and McMaster University, Hamilton, Canada.

## Author contributions

DW, MAR, CPV, and DB developed the study concept and design. DW and AGG performed laboratory analysis. DW, MAR, AG, MT, and CPV did data acquisition, analysis, and interpretation. DW drafted the manuscript. CPV performed bioinformatics analysis. All authors reviewed and endorsed the final version of the manuscript.

## Data availability

All data related to this study will be made available upon request and review.

## Supplemental materials

**Supplementary Figure 1.**
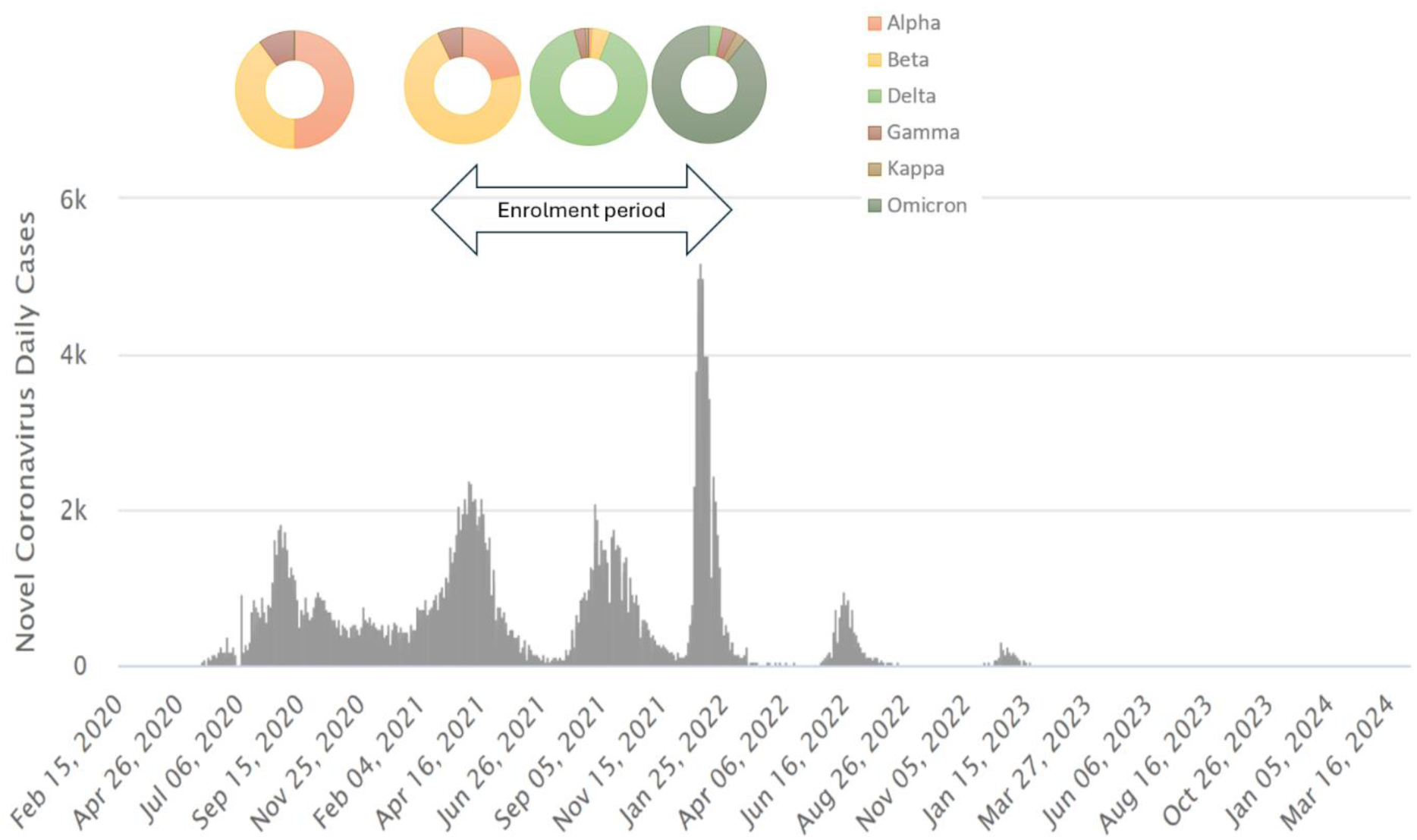
The four major COVID-19 waves in Ethiopia (**Source:** https://www.worldometers.info/coronavirus/country/ethiopia) and the corresponding SARS-CoV-2 variants circulating in the country during the different pandemic waves. Data for the SARS-CoV-2 genotyping is obtained from EPHI and has been published previously (44–46). The cohort for the current study enrolled between waves 2 and 4.

**Supplementary Figure 2.**
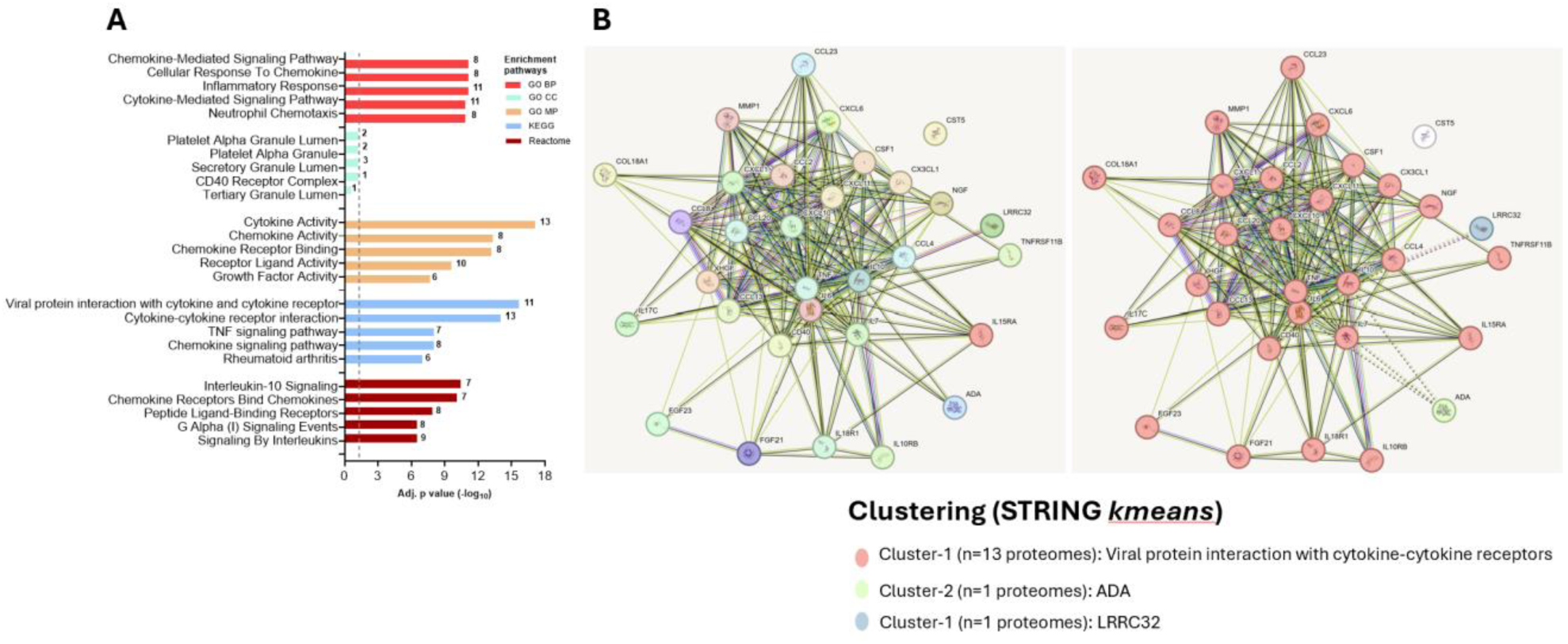
Gene Ontology (GO), KEGG and Reactome pathways (A) and STRING protein-protein interaction (B).

**Supplementary Figure 3.**
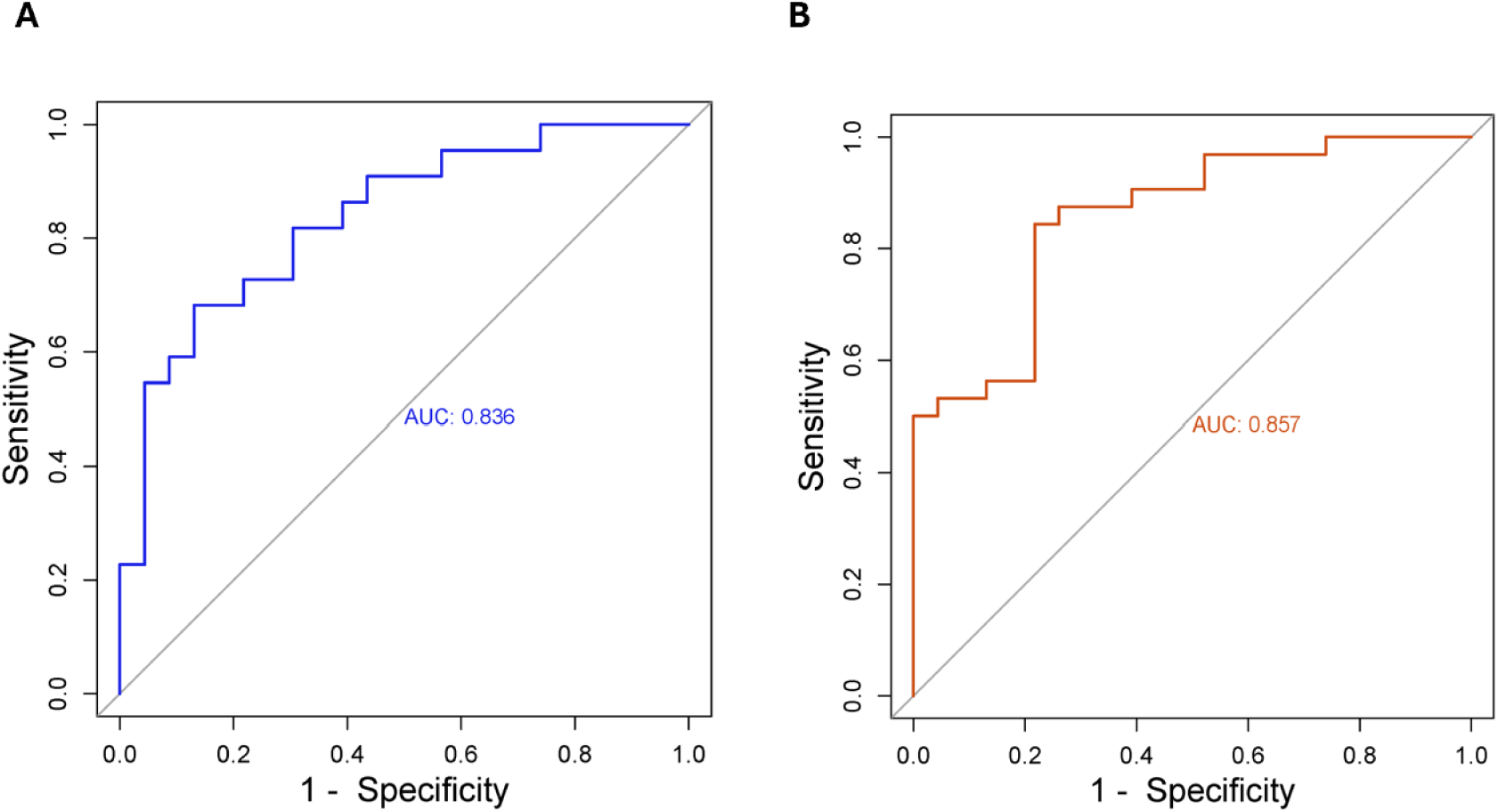
Combined AUC scores of SLAMF1, IL15RA, and IL18 in critical COVID-19 (A), or TNF, IL15RA and IL18 in Long COVID (B)

**Supplementary Table 1.** Baseline Clinical Characteristics of Study Participants.

| Characteristic <sup>a</sup> | All COVID-19<br>(n =67) | Non-critical COVID-19<br>(n =45) | Critical COVID-19<br>(n =22) | P value* |
| --- | --- | --- | --- | --- |
| Headache | 25 (45.5) | 11 (33.3) | 14 (63.6) | **0.027** |
| Loss of smell | 27 (49.1) | 12 (36.4) | 15 (68.2) | **0.021** |
| Loss of taste | 25 (45.5) | 11 (33.3) | 14 (63.6) | **0.027** |
| Cough | 22 (40.0) | 5 (15.2) | 17 (77.3) | **<0.0001** |
| Shortness of breath | 17 (30.9) | 4 (12.1) | 13 (59.1) | **<0.0001** |
| Wheezing | 13 (23.6) | 6 (18.2) | 7 (31.8) | 0.244 |
| Chest pain | 22 (40.0) | 9 (27.3) | 13 (59.1) | **0.018** |
| Sore throat | 26 (47.3) | 9 (27.3) | 17 (77.3) | **<0.0001** |
| Nasal congestion | 25 (45.5) | 13 (39.4) | 12 (54.6) | 0.269 |
| Anorexia | 25 (45.5) | 14 (42.2) | 11 (50.0) | 0.580 |
| Vomiting/nausea | 12 (21.8) | 11 (33.3) | 1 (4.6) | **0.011** |
| Diarrhea | 8 (14.6) | 7 (21.2) | 1 (4.6) | 0.086 |
| Abdominal pain | 9 (16.4) | 7 (21.2) | 2 (9.1) | 0.234 |
| Arthralgia | 29 (52.7) | 13 (39.4) | 16 (72.7) | **0.015** |
| Myalgia | 28 (50.9) | 12 (36.4) | 16 (72.7) | **0.008** |
| Fatigue or malaise | 30 (54.6) | 13 (39.4) | 17 (77.3) | **0.006** |
| Fever | 28 (50.9) | 12 (36.4) | 16 (72.7) | **0.008** |
| Conjunctivitis | 18 (32.7) | 15 (45.5) | 3 (13.6) | **0.014** |
| Pneumonia | 15 (27.3) | 0 (0.0) | 15 (68.2) | **<0.0001** |

<sup>a</sup>Data is number (%). \*Differences calculated using Pearson's Chi2 or Fisher's exact test where appropriate.

**Supplementary Table 2.** Inflammatory panel proteomics assessed in this study.

|  |  |
| --- | --- |
| ADA | Adenosine Deaminase |
| ARTN | Artemin |
| AXIN1 | Axin-1 |
| Beta-NGF | Beta-nerve growth factor |
| CASP-8 | Caspase-8 |
| CCL9 | C-C motif chemokine 19 |
| CCL20 | C-C motif chemokine 20 |
| CCL23 | C-C motif chemokine 23 |
| CCL25 | C-C motif chemokine 25 |
| CCL28 | C-C motif chemokine 28 |
| CCL3 | C-C motif chemokine 3 |
| CCL4 | C-C motif chemokine 4 |
| CD40 | CD40L receptor |
| CDCP1 | CUB domain-containing protein 1 |
| CXCL1 | C-X-C motif chemokine 1 |
| CXCL10 | C-X-C motif chemokine 10 |
| CXCL11 | C-X-C motif chemokine 11 |
| CXCL5 | C-X-C motif chemokine 5 |
| CXCL6 | C-X-C motif chemokine 6 |
| CXCL9 | C-X-C motif chemokine 9 |
| CST5 | Cystatin D |
| DNER | Delta and Notch-like epidermal growth factor-related receptor |
| CCL11 | Eotaxin |
| 4E-BP1 | Eukaryotic translation initiation factor 4E-binding protein 1 |
| FGF-19 | Fibroblast growth factor 19 |
| FGF-21 | Fibroblast growth factor 21 |
| FGF-23 | Fibroblast growth factor 23 |
| FGF-5 | Fibroblast growth factor 5 |
| Flt3L | Fms-related tyrosine kinase 3 ligand |
| CX3CL1 | Fractalkine |
| GDNF | Glial cell line-derived NEUtrophic factor |
| HGF | Hepatocyte growth factor |
| IFN-gamma | Interferon gamma |
| IL-1 alpha | Interleukin-1 alpha |
| IL10 | Interleukin-10 |
| IL-10RA | Interleukin-10 receptor subunit alpha |
| IL-10RB | Interleukin-10 receptor subunit beta |
| IL-12B | Interleukin-12 subunit beta |
| IL-13 | Interleukin-13 |
| IL-15RA | Interleukin-15 receptor subunit alpha |
| IL-17A | Interleukin-17A |
| IL-17C | Interleukin-17C |
| IL-18 | Interleukin-18 |
| IL-18R1 | Interleukin-18 receptor 1 |
| IL-2 | Interleukin-2 |
| IL-2RB | Interleukin-2 receptor subunit beta |
| IL-20 | Interleukin-20 |
| IL-20RA | Interleukin-20 receptor subunit alpha |
| IL-22 RA1 | Interleukin-22 receptor subunit alpha-1 |
| IL-24 | Interleukin-24 |
| IL-33 | Interleukin-33 |
| IL-4 | Interleukin-4 |
| IL5 | Interleukin-5 |
| IL6 | Interleukin-6 |
| IL-7 | Interleukin-7 |
| IL-8 | Interleukin-8 |
| LAP TGF-beta-1 | Latency-associated peptide transforming growth factor beta-1 |
| LIF | Leukemia inhibitory factor |
| LIF-R | Leukemia inhibitory factor receptor |
| CSF-1 | Macrophage colony-stimulating factor 1 |
| MMP-1 | Matrix metalloproteinase-1 |
| MMP-10 | Matrix metalloproteinase-10 |
| MCP-1 | Monocyte chemotactic protein 1 |
| MCP-2 | Monocyte chemotactic protein 2 |
| MCP-3 | Monocyte chemotactic protein 3 |
| MCP-4 | Monocyte chemotactic protein 4 |
| CD244 | Natural killer cell receptor 2B4 |
| NT-3 | NEUtrophin-3 |
| NRTN | Neurturin |
| OSM | Oncostatin-M |
| OPG | Osteoprotegerin |
| PD-L1 | Programmed cell death 1 ligand 1 |
| EN-RAGE | Protein S100-A12 |
| SLAMF1 | Signaling lymphocytic activation molecule |
| SIRT2 | SIR2-like protein 2 |
| STAMPB | STAM-binding protein |
| SCF | Stem cell factor |
| ST1A1 | Sulfotransferase 1A1 |
| CD6 | T cell surface glycoprotein CD6 isoform |
| CD5 | T-cell surface glycoprotein CD5 |
| CD8A | T-cell surface glycoprotein CD8 alpha chain |
| TSLP | Thymic stromal lymphopoietin |
| TNFB | TNF-beta |
| TRANCE | TNF-related activation-induced cytokine |
| TRAIL | TNF-related apoptosis-inducing ligand |
| TGF-alpha | Transforming growth factor alpha |
| TWEAK | Tumor necrosis factor Ligand superfamily member 12 |
| TNF | Tumor necrosis factor |
| TNFSF14 | Tumor necrosis factor ligand superfamily member 14 |
| TNFRSF9 | Tumor necrosis factor receptor superfamily member 9 |
| uPA | Urokinase-type plasminogen activator |
| VEGF-A | Vascular endothelial growth factor A |

